# Serological Cross-Reactivity Between Spotted Fever and Typhus Groups of Rickettsia Infection in Japan

**DOI:** 10.1101/2022.10.16.22281137

**Authors:** Tetsuro Aita, Eiichiro Sando, Shungo Katoh, Sugihiro Hamaguchi, Hiromi Fujita, Noriaki Kurita

## Abstract

We examined the frequency of cross-reactions to *Rickettsia typhi* in patients with Japanese spotted fever and found that approximately 20% of cases were positive. Comparison of antibody titers revealed the difficulty in identifying some positive cases. The frequency of misclassification in serodiagnosis was found to be limited.

## [Introduction]

Conventional serodiagnosis using indirect immunofluorescence assay (IFA) or indirect immunoperoxidase assay (IP) in paired sera remains a cornerstone for acute infections of two antigenic groups of the genus *Rickettsia*: the spotted fever group (SFG) and typhus group (TG) (1). However, serological cross-reactions between different groups can occur and cause misclassification during serodiagnosis, leading to misdiagnosis, especially in patients with non-specific clinical features (1). The cross-reactivity within SFG was previously investigated (2), whereas that between SFG and TG is yet to be fully evaluated. Uchiyama et al. identified cross-reactive antibodies of *Rickettsia typhi*, a TG rickettsia causing murine typhus (MT), with those of *Rickettsia japonica*, an SFG rickettsia causing Japanese spotted fever (JSF), in patients with JSF using western immunoblotting and cross-absorption (3). However, these assays are difficult to implement as standard diagnostic tests because they require expertise. Clinicians must speculate on cross-reactions based on clinical features and conventional serological results in paired sera. Although experts have proposed that the variation of each rickettsial antibody titer in two phases differentiates a true infection from a cross-reaction (4), no clinical evidence is available to support this. Therefore, using a conventional method, we aimed to investigate the cross-reactions between *R. japonica* and *R. typhi* in paired sera from patients with clinically and serologically confirmed *R. japonica* infections.

## [Methods]

We retrospectively examined serological data from two reference centers for rickettsiosis in Japan between January 1, 2003, and December 31, 2016. All patient samples were tested by IP using the Aoki strain of *R. japonica* and the Wilmington strain of *R. typhi* (Appendix). In the present study, patients with JSF were defined as having clinical features of the disease and a ≥4-fold increase in IgM or IgG titers against *R. japonica* or seroconverted (defined as titers ≥80) in paired sera (5).

The cross-reaction was defined as a higher titer against *R. typhi* in convalescent sera than that in acute sera (2), and its frequency was evaluated for IgM and IgG. We also investigated the presence of cross-reactivity in patients diagnosed with JSF using polymerase chain reaction (PCR) or immunohistochemistry in skin biopsy specimens (6). Additionally, the patient samples that indicated cross-reactivity but posed challenges in being identified as either JSF or MT based on IP alone, were subjected to indirect hemagglutination tests (HA) (7,8).

## [Results]

A total of 145 patients diagnosed with JSF were analyzed. The median interval between acute and convalescent serum collection was 13 (interquartile range, 8–17) days. The changes in titers for *R. japonica* and *R. typhi* are indicated in the Appendix Figure. The cross-reactions against *R. typhi* in patients with JSF were observed in 21.4% of cases (27/126, 95% confidence interval [CI] 14.6–29.6) for IgM and 19.6% (27/138, 95% CI 13.3–27.2) for IgG (Figure). IgM and IgG titers are indicated in Appendix Table 1 and Table, respectively. Additionally, in 11 cases of JSF that were confirmed using PCR or immunohistological tests, cross-reactions were observed in 18.2% of cases (2/11, 95% CI 2.3–51.8) each for IgM and IgG, with the titers listed in Appendix Table 2. Eight cases could not be identified as true infections or cross-reactions by comparing endpoint titer levels of two rickettsiae; seven cases of these could be differentiated using HA (Appendix Tables 3 and 4).

**Figure.**
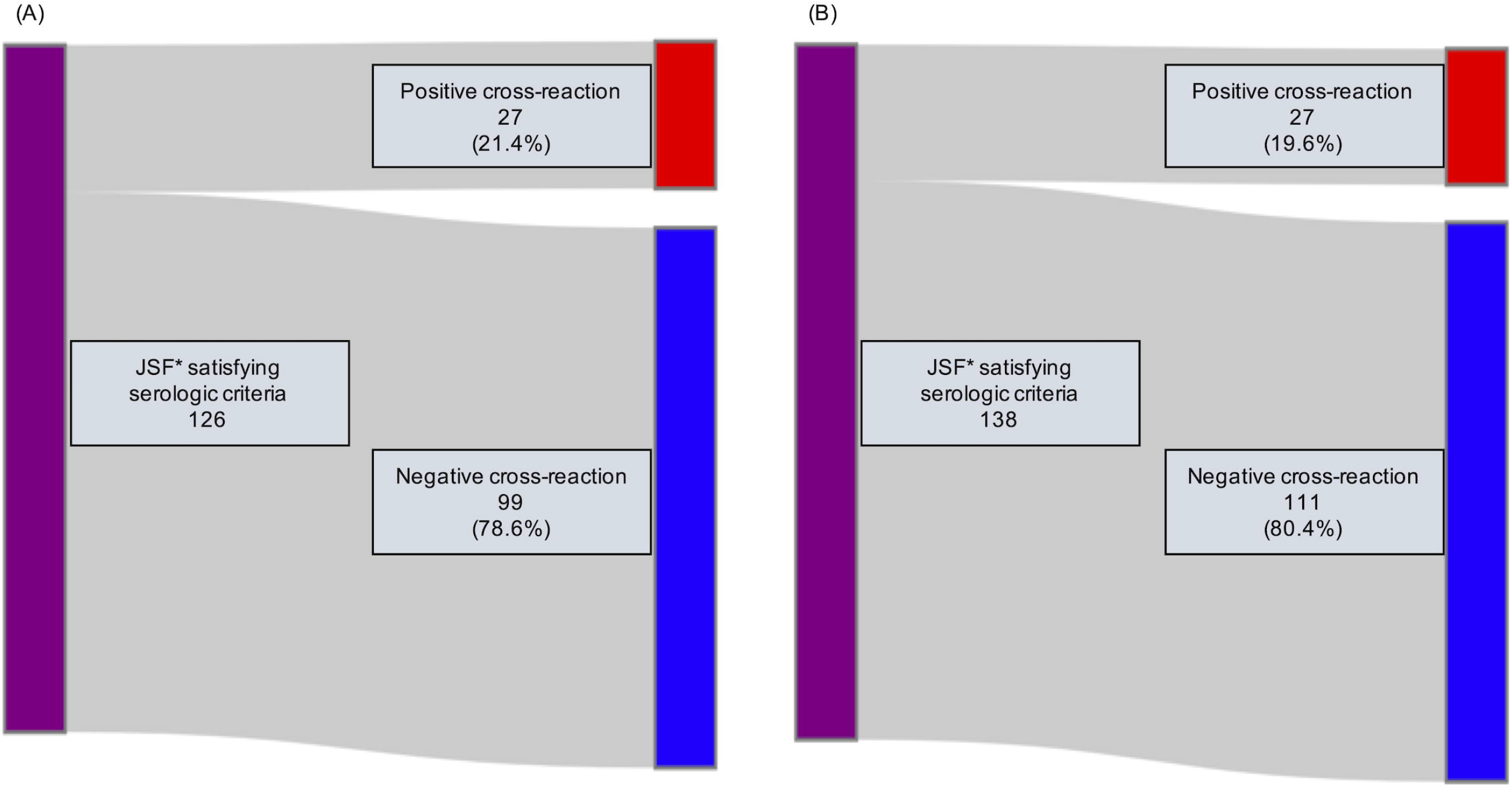
The cross-reactions against *Rickettsia typhi* in patients with Japanese spotted fever (A) IgM (B) IgG *JSF, Japanese spotted fever Of the 145 cases analyzed, 119 underwent a ≥4-fold increase in titers against *R. japonica* or the occurrence of seroconversion in paired sera both for IgM and IgG, with 7 and 19 having these serological reactions only in IgM and IgG, respectively (total number of analyzed cases with IgM, 126; those with IgG, 138).

**Table.**
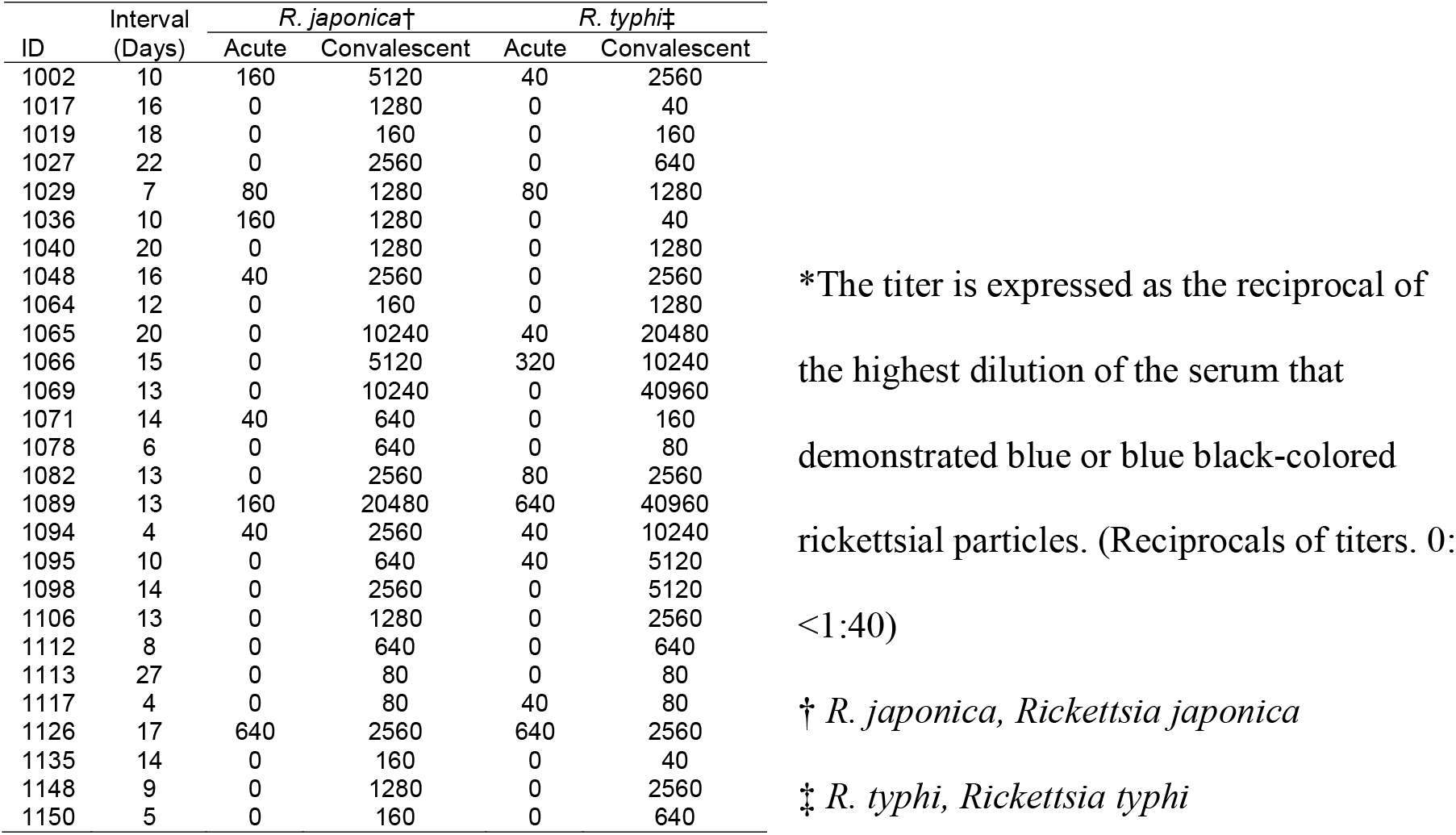
The antibody titers against *Rickettsia japonica* and *R. typhi* in patients exhibiting cross-reactions in IgG*

## [Discussion]

In this study, using conventional serological methods in paired sera, we identified that the proportion of cross-reactions between *R. japonica* and *R. typhi* in patients with JSF was approximately 20%, suggesting a limited possibility of misclassification in serodiagnosis. The robustness of our results was supported by a similar frequency of cross-reactions in patients with JSF that was confirmed using PCR or immunohistological skin findings.

To our knowledge, only one study has investigated the serological cross-reactions between JSF and MT. A previous study identified cross-reactions between *R. japonica* and *R. typhi* in patients with JSF using IFA, immunoblotting, and cross-adsorption assays, focusing on the identification of proteins with common antigenicity using a small sample size (3). Thus, our study is the first to estimate the frequency of cross-reactions using a large number of patients with JSF and evaluate distinguishability using changes in antibody titers.

The value of endpoint antibody titers was proposed to differentiate SFG from TG (2,4,9), but this remains controversial. In fact, using a case definition of clinically and serologically confirmed (IP) patients with JSF, some patients with JSF with cross-reactions had high endpoint titers of *R. typhi*.

Especially in cases with non-specific rickettsial symptoms, the cross-reaction results in our study may provide other possibilities: co-infection with *R. japonica* and *R. typhi*, cross-reactions against *R. typhi* in patients with *R. japonica* infection and previous *R. typhi* infection, or even cross-reactions against *R. japonica* in patients with true *R. typhi* infection.

## Supporting information

STROBE

## Data Availability

All data produced in the present work are contained in the manuscript.

## Acknowledgments

This work was supported by JSPS KAKENHI (Grant Number JP 19K23972) and AMED (Grant Number JP21fk0108614).

## Author Bio

T. Aita is an internist specializing in general internal medicine and clinical epidemiology. He is affiliated with the Department of General Internal Medicine, Fukushima Medical University as a teaching/research associate and is interested in research on the epidemiology of diseases, diagnostic accuracy, and infections, such as rickettsia.

## Appendix

### Indirect immunoperoxidase assay used in this study

The IP assays were implemented at Ohara Research Laboratory, Ohara General Hospital in Fukushima, and Mahara Institute of Medical Acarology in Tokushima.

The rickettsial strains were grown in L929 cells and used as antigens. The antigen was suspended in 0.01 M phosphate-buffered saline (PBS) with 0.3% bovine serum albumin fraction V at pH 7.2. The antigen was spotted on a slide, diluted to 100–200 rickettsial particles per field of a microscope (×40), air-dried for 30 min, and fixed in acetone for 10 min at room temperature. The slide-antigen was immediately utilized after drying, although it was maintained at −20 °C. Patient sera were diluted 2-fold in PBS, from 1:40 to 1:40960, and 0.01 mL of each dilution was placed on the antigen spot on the slide, incubated for 30 min at 36 °C in a humidified room, and washed twice in PBS for 5 min. Subsequently, each spot received 0.01 mL of 1:100 diluted peroxidase-conjugated antihuman IgG or IgM rabbit serum (γ-chain specific or μ-chain specific; DAKO Corp., Glostrup, Denmark), incubated for 30 min at 36 °C in the chamber, and rinsed as indicated above. Finally, the slide was placed in a freshly produced enzyme substrate solution comprising 1 volume of 80% ethanol containing 0.2% 4-Cl-1-naphthol, 4 volumes of PBS, and volume of 3% peroxide for 5 min at room temperature. This was washed twice in distilled water for 5 min each time and then dried. The slide was then sealed with a coverslip and glycerol gelatin. A microscope was used to visually read the results. The titer was calculated as the reciprocal of the highest serum dilution that showed blue or blue-black rickettsial particles.

### Appendix Tables

**Table 1.** The antibody titers against *Rickettsia japonica* and *R. typhi* in patients with the cross-reactions in IgM*

| ID | Interval<br>(Days) | *R. japonica*† |  | *R. typhi*‡ |  |
| --- | --- | --- | --- | --- | --- |
|  |  | Acute | Convalescent | Acute | Convalescent |
| 1005 | 8 | 80 | 320 | 0 | 40 |
| 1007 | 14 | 0 | 1280 | 80 | 640 |
| 1017 | 16 | 0 | 160 | 0 | 40 |
| 1019 | 18 | 0 | 640 | 0 | 40 |
| 1020 | 10 | 0 | 320 | 0 | 160 |
| 1027 | 22 | 0 | 320 | 0 | 160 |
| 1033 | 15 | 0 | 320 | 0 | 40 |
| 1036 | 10 | 0 | 2560 | 0 | 80 |
| 1041 | 18 | 0 | 640 | 0 | 160 |
| 1044 | 14 | 0 | 10240 | 0 | 40 |
| 1048 | 16 | 0 | 640 | 0 | 40 |
| 1053 | – | 0 | 10240 | 0 | 40 |
| 1064 | 12 | 0 | 640 | 0 | 40 |
| 1069 | 13 | 0 | 160 | 0 | 40 |
| 1077 | 13 | 0 | 640 | 0 | 40 |
| 1078 | 6 | 0 | 160 | 0 | 40 |
| 1081 | 8 | 0 | 5120 | 0 | 80 |
| 1082 | 13 | 0 | 2560 | 0 | 160 |
| 1085 | 23 | 0 | 2560 | 0 | 40 |
| 1089 | 13 | 80 | 640 | 160 | 1280 |
| 1093 | 7 | 0 | 1280 | 0 | 80 |
| 1094 | 4 | 0 | 640 | 0 | 640 |
| 1096 | 7 | 0 | 320 | 0 | 160 |
| 1097 | 7 | 0 | 320 | 0 | 160 |
| 1118 | 13 | 0 | 640 | 0 | 80 |
| 1147 | 13 | 0 | 5120 | 0 | 80 |
| 1150 | 5 | 0 | 160 | 0 | 320 |

\*The titer is expressed as the reciprocal of the highest dilution of the serum that demonstrated blue or blue black-colored rickettsial particles. (Reciprocals of titers, 0: <1:40) –, No data.
† *R. japonica*, *Rickettsia japonica*
‡ *R. typhi*, *Rickettsia typhi*

**Table 2.** The antibody titers against *Rickettsia japonica* and *R. typhi* in patients with Japanese spotted fever confirmed using polymerase chain reaction or immunohistological tests*

(A)IgM
| ID | Interval<br>(Days) | *R. japonica*† |  | *R. typhi*‡ |  |
| --- | --- | --- | --- | --- | --- |
|  |  | Acute | Convalescent | Acute | Convalescent |
| 1018 | 29 | 0 | 160 | 0 | 0 |
| 1019 | 18 | 0 | 640 | 0 | 40 |
| 1023 | 21 | 0 | 640 | 0 | 0 |
| 1027 | 22 | 0 | 320 | 0 | 160 |
| 1031 | 12 | 0 | 1280 | 0 | 0 |
| 1037 | 15 | 0 | 160 | 0 | 0 |
| 1050 | 16 | 0 | 4960 | 0 | 0 |
| 1057 | 29 | 0 | 320 | 0 | 0 |
| 1070 | 15 | 0 | 640 | 0 | 0 |
| 1141 | 13 | 0 | 320 | 0 | 0 |
| 1142 | 10 | 0 | 320 | 0 | 0 |

(B)IgG
| ID | Interval<br>(Days) | *R. japonica*† |  | *R. typhi*‡ |  |
| --- | --- | --- | --- | --- | --- |
|  |  | Acute | Convalescent | Acute | Convalescent |
| 1018 | 29 | 0 | 320 | 0 | 0 |
| 1019 | 18 | 0 | 160 | 0 | 160 |
| 1023 | 21 | 0 | 320 | 0 | 0 |
| 1025 | 21 | 0 | 640 | 0 | 0 |
| 1027 | 22 | 0 | 2560 | 0 | 640 |
| 1031 | 12 | 0 | 640 | 0 | 0 |
| 1037 | 15 | 160 | 10240 | 0 | 0 |
| 1049 | 13 | 0 | 640 | 0 | 0 |
| 1050 | 16 | 0 | 2560 | 0 | 0 |
| 1057 | 29 | 0 | 640 | 0 | 0 |
| 1070 | 15 | 0 | 5120 | 0 | 0 |

\*The titer is expressed as the reciprocal of the highest dilution of the serum that demonstrated blue or blue black-colored rickettsial particles. (Reciprocals of titers, 0: <1:40)
† *R. japonica*, *Rickettsia japonica*
‡ *R. typhi*, *Rickettsia typhi*

**Table 3.**
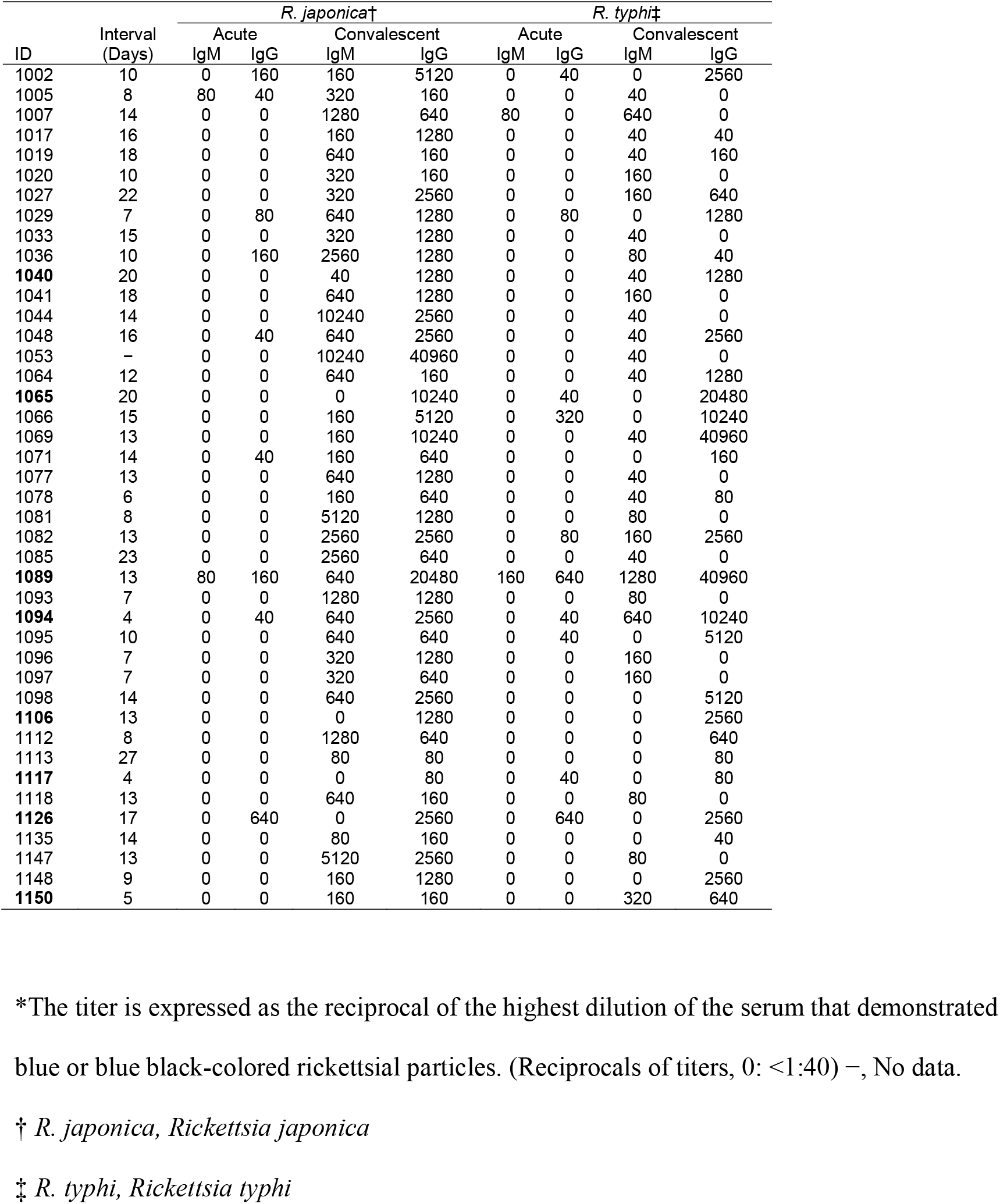
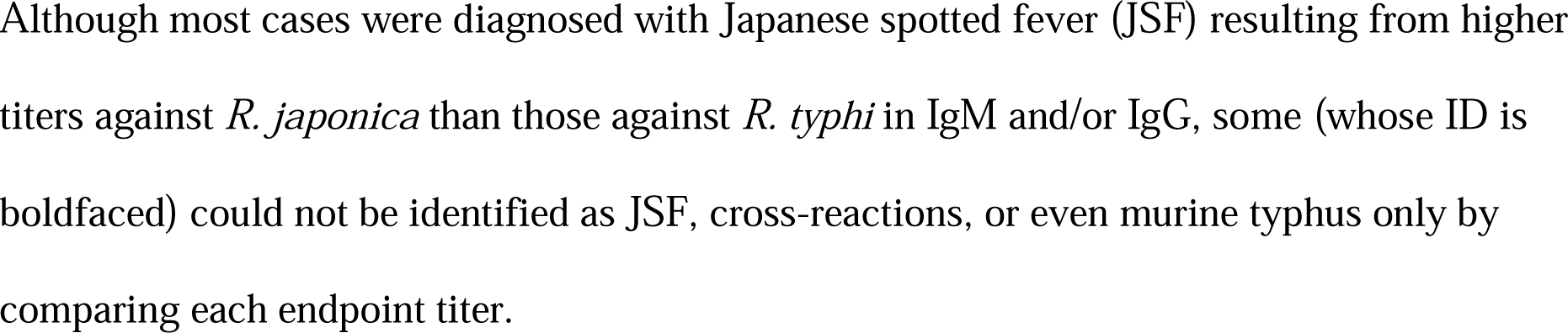
All the antibody titers against *Rickettsia japonica* and *R. typhi* in patients with Japanese spotted fever who exhibited cross-reactions*

**Table 4.**
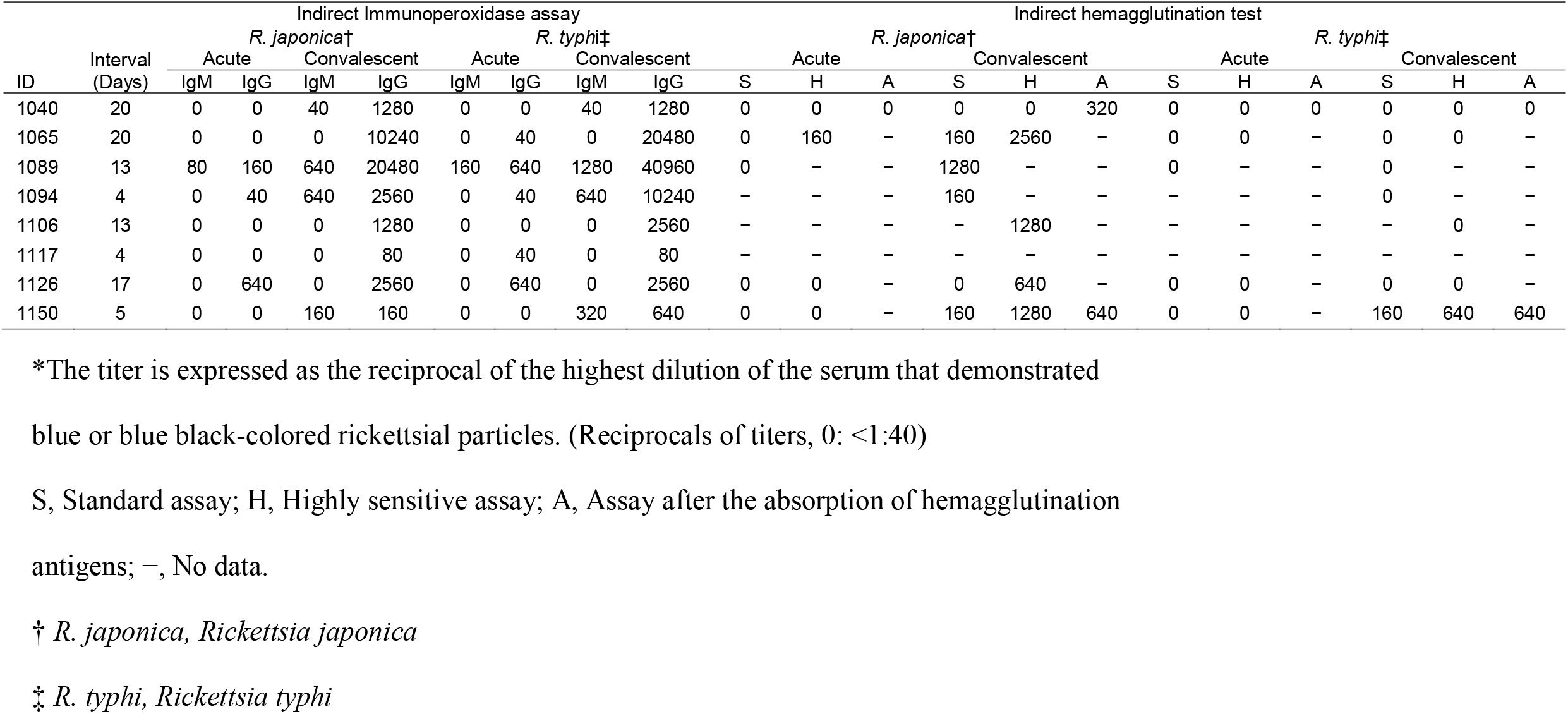
The antibody titers obtained using the indirect hemagglutination test against *Rickettsia japonica* and *R. typhi* in patients with Japanese spotted fever (JSF) who exhibited cross-reactions but whose disease could not be identified as JSF or Murine typhus based only on the result of the indirect immunoperoxidase assay*

### Appendix Figure

**Figure.**
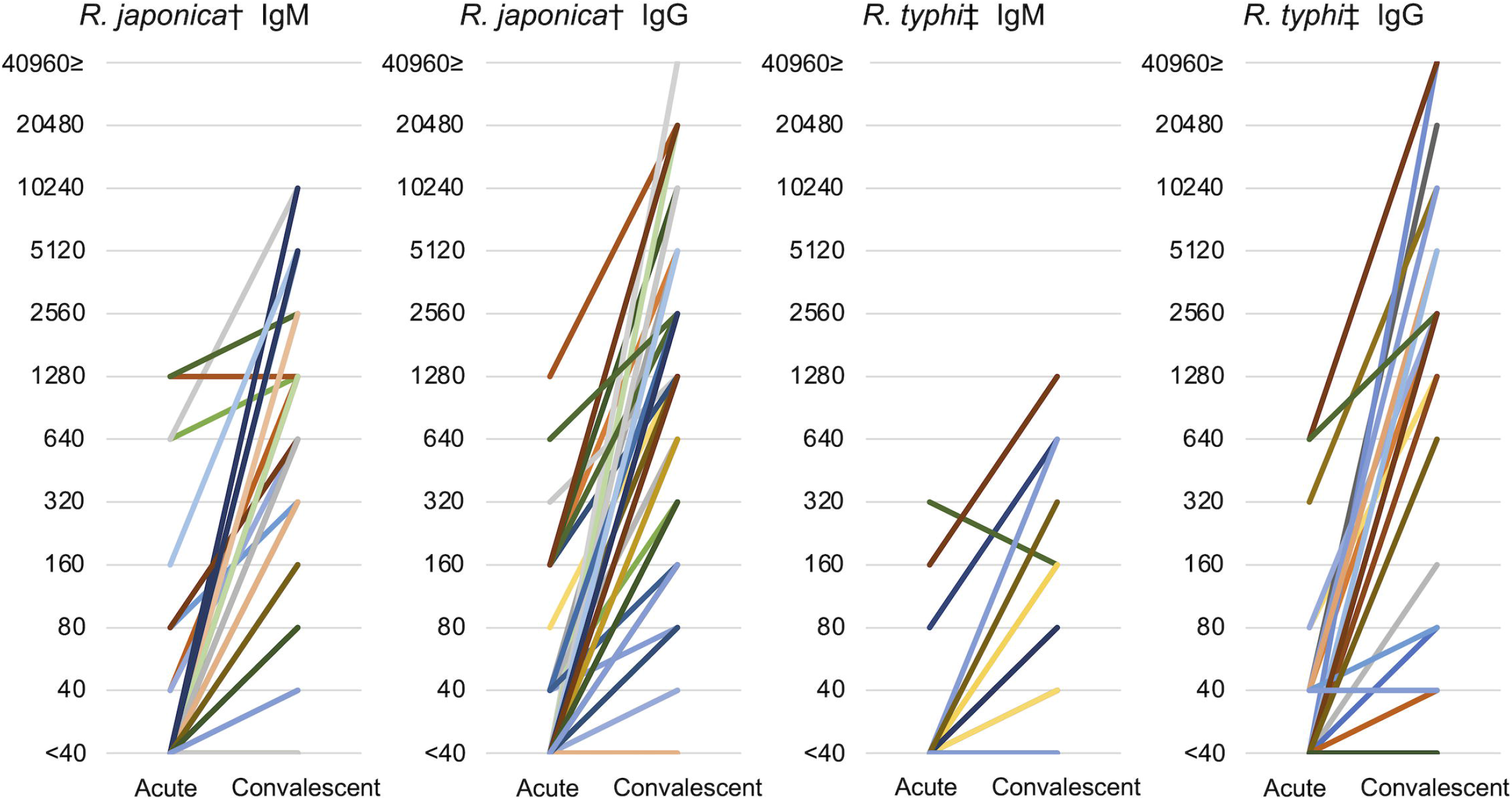
The change in titers for *Rickettsia japonica* and *R. typhi* between acute and convalescent sera* *The titer is expressed as the reciprocal of the highest dilution of the serum that demonstrated blue or blue black-colored rickettsial particles. † *R. japonica, Rickettsia japonica* ‡ *R. typhi, Rickettsia typhi*

## Notes

### Competing Interest Statement

The authors have declared no competing interest.

### Author Declarations

The Institutional Review Board of Fukushima Medical University gave ethical approval for this work.

